# Thyroid function and amyotrophic lateral sclerosis: a mendelian randomization study

**DOI:** 10.1101/2025.05.21.25328118

**Authors:** Ezekiel Damilare Jacobs, Adeyemi Timothy Akinade, Chucks Marvellous Obere

## Abstract

**Background:** Amyotrophic lateral sclerosis (ALS) is a rare, progressive neurodegenerative disorder causing motor neuron degeneration, muscle paralysis, and death within 3–5 years, with a rising global prevalence. While thyroid dysfunction is implicated in the pathology of other neurodegenerative diseases, its role in ALS remains unclear due to conflicting reports from observational studies. To address this, we aimed to investigate the causal relationship between thyroid function and ALS using two-sample Mendelian Randomization (MR).

**Method:** We performed a two-sample MR study to evaluate the bidirectional causal relationship between thyroid function traits (TSH, FT4, autoimmune hyperthyroidism, autoimmune hypothyroidism) and ALS. We used SNPs from GWAS data (TSH and FT4: n=271,040; autoimmune hyperthyroidism: n=1,828 cases/279,855 controls; autoimmune hypothyroidism: n=40,926 cases/274,069 controls; ALS: n=27,205 cases/110,881 controls). We applied the inverse-variance weighted (IVW) method as the primary analysis, with sensitivity analyses (MR-Egger, weighted median, weighted mode, MR-PRESSO) to assess pleiotropy and heterogeneity.

**Results:** We identified a causal effect of autoimmune hyperthyroidism on reducing ALS risk (IVW OR = 0.96; 95% CI: 0.93–0.99; P = 0.03), but no associations for TSH (IVW OR = 0.99; 95% CI: 0.94–1.04; P = 0.71), FT4 (IVW OR = 1.03; 95% CI: 0.96–1.12; P = 0.36), or autoimmune hypothyroidism (IVW OR = 1.01; 95% CI: 0.98–1.05; P = 0.39) with ALS. Bidirectional analysis of genetic liability to ALS showed no causal effect on TSH (IVW OR = 0.98; 95% CI: 0.96–1.01; P = 0.65), FT4 (IVW OR = 0.98; 95% CI: 0.94–1.02; P = 0.46), autoimmune hypothyroidism (IVW OR = 0.93; 95% CI: 0.76–1.13; P = 0.59), or autoimmune hyperthyroidism (IVW OR = 1.01; 95% CI: 0.76–1.33; P = 0.78).

**Conclusion:** We found that genetic predisposition to autoimmune hyperthyroidism may reduce ALS risk. Our findings expand the knowledge of its genetic underpinnings and lay a foundation for future research into therapeutic strategies targeting thyroid function to mitigate ALS risk.

## INTRODUCTION

Amyotrophic lateral sclerosis (ALS) is a rare, progressive neurodegenerative disease that affects motor neurons and brain cells that regulate voluntary movement and breathing (1,2). This condition leads to worsening symptoms such as muscle paralysis and atrophy, respiratory failure and eventually death usually occurring within 3-5 years from onset of symptoms (2–4).

The global burden of ALS remains unclear due to the disease’s rarity and variations in incidence and prevalence across regions (2). Published reports indicate that the global prevalence of ALS is projected to rise at an average annual rate of 2.12% through 2040, with estimates increasing from approximately 3.1 to 5.2 cases per 100,000 people between 2015 and 2040 (4–10). Amyotrophic lateral sclerosis (ALS) affects all racial groups, although most studies indicate a higher prevalence among white and non-Hispanic populations (5). However, a recent review highlighted that ALS may be more prevalent in Africa than in developed countries and associated with an even worse prognosis, attributing this underrepresentation of facts and figures to insufficient data and perhaps ALS diagnostic challenges, which often take up to 14 months (8,11). In the coming decades, the increasing prevalence of ALS is expected to shift toward developing countries due to the ageing population in those regions (5).

The age group that is mostly at risk is between the ages of 50 and 70 (2,4). However, recently, a variation of ALS that affects children and other young persons has been reported (12,13). Also, the male sex is cited as a greater risk factor than the female sex, with early testosterone influence implicated in its pathogenesis (9,11). About 85- 90% of ALS is sporadic (i.e. not hereditary), whereas around 10% is hereditary (familial). Genetic risk factors such as mutations in the superoxide dismutase 1 (*SOD1*) and chromosome 9 open reading frame 72 (*C9orf72*) genes have been associated with both sporadic and familial forms, with a stronger association with familial ALS (4,14). Environmental factors such as air pollution, organic dust, and exposure to heavy metals, have also been associated with sporadic ALS. Additionally, lifestyle factors including smoking, physical activity, diet, military service, and traumatic brain injury, have been suggested as potential risk factors, although definitive conclusions are lacking (4,8,11). Currently, there is no known treatment that stops or reverses the progression of ALS. However, some combination of therapies may slow progression of the disease, improve quality of life, and extend survival.

Thyroid function and its dysfunction has been implicated in several neurodegenerative disorders, including Alzheimer’s disease, multiple sclerosis, and Parkinson’s disease (15–18). However, the relationship between thyroid function and amyotrophic lateral sclerosis (ALS) remains poorly defined. To date, few observational studies have investigated the association between thyroid function and ALS. With mixed findings, some suggest a potential link with thyroid abnormalities, while others report no significant association (19–21). For instance, a large-scale retrospective study which used propensity-score matching reported an increased risk of ALS associated with thyroid disorders (20). In contrast, another case-control study of ALS patients found no meaningful differences in thyroid function between ALS patients and controls (21). These inconsistencies may be attributed to differences in study design, population heterogeneity, or unmeasured confounding, and thereby highlight the need for more efficient approaches that can strengthen causal inference.

To address this, we have used a causal inference approach (Mendelian Randomisation) that minimise biases such as residual confounding and measurement errors commonly associated with conventional observational studies (22,23). Mendelian Randomisation (MR) uses genetic variants such as single nucleotide polymorphisms (SNPs) strongly associated with the exposure of interest (23). These genetic variants are then used as instrumental variables to evaluate the causal effect of genetic predisposition to the exposure on the outcome (23). In this study, we aimed to investigate whether there is a causal relationship between genetically predicted thyroid function traits and Amyotrophic lateral sclerosis, using a two-sample MR.

## METHODS

### Study Design

This study employed a two-sample Mendelian Randomization (MR) approach, which is an extension of the traditional MR that typically relies on single-sample data. The two-sample MR differs primarily in its use of genome-wide association studies (GWAS) data from independent samples within the same underlying population to evaluate the causal relationship between an exposure and an outcome (24). This approach enhances statistical power and minimizes biases associated with overlapping samples, and thus allows for a more robust causal inference (23,24). The reliability of the conclusions drawn through MR is predicated upon three core assumptions that the genetic instruments used must satisfy, as summarized in Figure 1.

**Figure 1:**
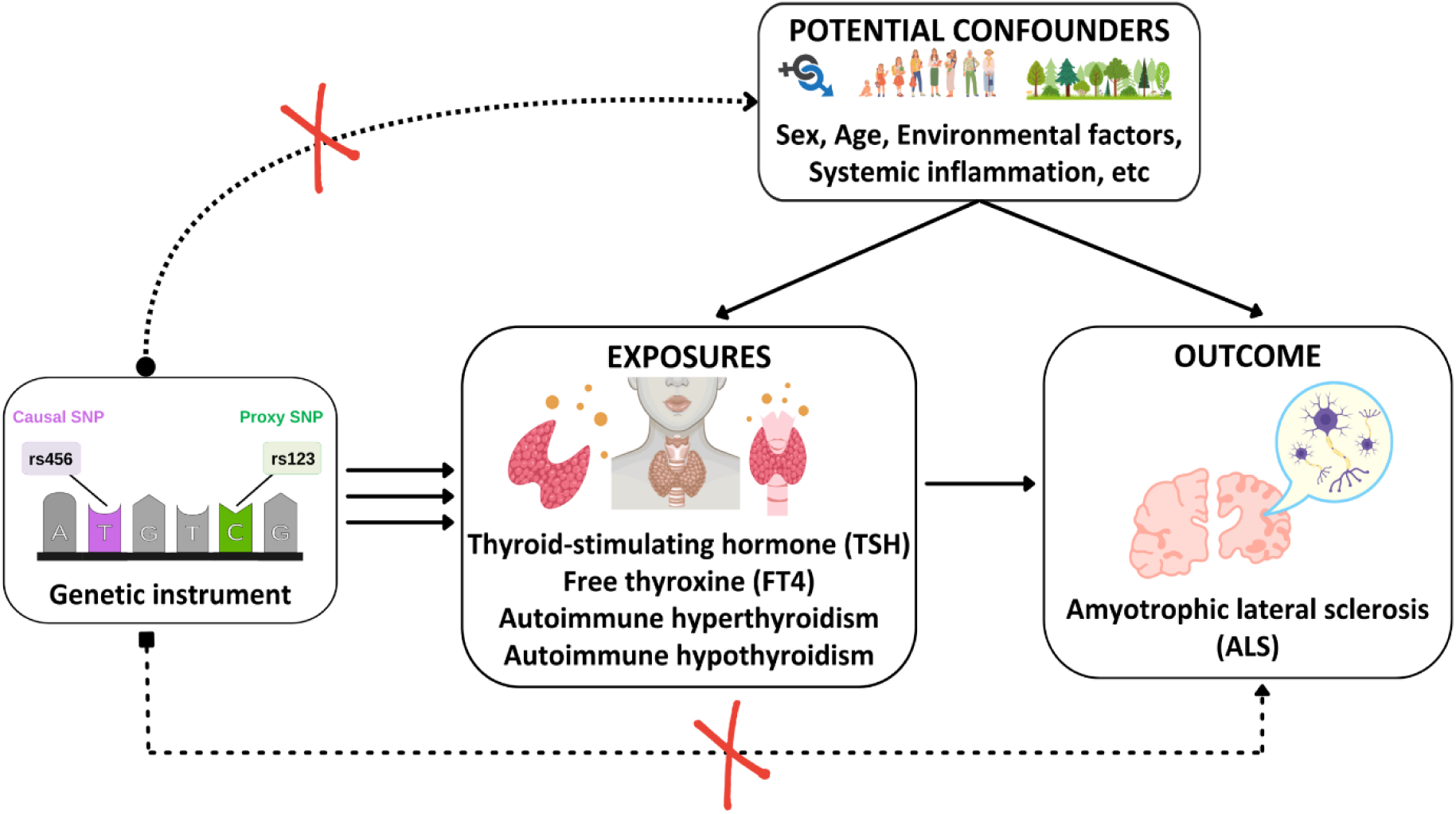
Schematic representation of the three core assumptions of Mendelian Randomization (MR) as adapted to this study (23,24): (1) The genetic instruments (SNPs) are strongly associated with the exposure. (2) The genetic instruments are independent of the confounders of the exposure–outcome relationship. (3) The genetic instruments affect the outcome only via their effect on the exposure, i.e. no horizontal pleiotropy.

### Data sources

We obtained genome-wide association study (GWAS) summary-level data for thyroid function traits, specifically normal range thyroid-stimulating hormone (TSH) and free thyroxine (FT4) levels, from a meta-analysis conducted by the ThyroidOmics Consortium, comprising 271,040 individuals. This analysis identified 259 loci associated with TSH levels and 85 genetic variants associated with FT4 levels (25).

Additional GWAS summary statistics were obtained from the FinnGen Release 9 dataset. The total sample comprised 244,737 participants, including individuals from the UK Biobank (UKB) and the Estonian Biobank (EstBB). For autoimmune hypothyroidism, the datasets comprised 40,926 cases and 274,069 controls. For autoimmune hyperthyroidism, there were 1,828 cases and 279,855 controls (26).

GWAS summary statistics for amyotrophic lateral sclerosis (ALS) were obtained from a large-scale meta-analysis conducted by van Rheenen et al. (2021) (27). The study included 27,205 cases and 110,881 controls, with 15 genome-wide significant loci identified.

As this study exclusively used publicly available, de-identified summary statistics from previously published GWAS, ethical approval and informed consent were not required.

Detailed information on these datasets is provided in Table 1

**Table 1:** Summary of the GWASs used in the two-sample MR analysis.

| Phenotype | PubMed ID | Sample/Cohort | Ancestry | Sample size |
| --- | --- | --- | --- | --- |
| Thyroid-stimulating hormone (TSH) | Sterenberg *et al.* (25) | ThyroidOmics Consortium | European (100%) | 271,040 |
| Free thyroxine (FT4) | Sterenberg *et al.* (25) | ThyroidOmics Consortium | European (100%) | 271,040 |
| Autoimmune hypothyroidism<br>(OpenGWAS ID: finn-b-E4_HYTHY_AI_STRICT) | Kurki *et al.* (26) | FinnGen R9, UK Biobank, Estonian Biobank | European (100%) | 40,926 cases<br>274,069 controls |
| Autoimmune hyperthyroidism<br>(Open GWAS ID: finn-b-AUTOIMMUNE_HYPERTHYROIDISM) | Kurki *et al.* (26) | FinnGen R9, UK Biobank, Estonian Biobank | European (100%) | 1,828 cases<br>279,855 controls |
| Amyotrophic lateral sclerosis (ALS)<br>(Open GWAS ID: ebi-a-GCST90027164) | van Rheenen *et al.* (27)<br>34873335 | MND Bank, QSkin, BePS, SLALOM, PARALS, SLAP, SLAGEN | European (100%) | 27,205 cases<br>110,881 controls |

### Genetic instrument extraction

We extracted independent SNPs at a genome-wide significance threshold (P < 5×10⁻⁸), applying linkage disequilibrium clumping (r² < 0.01; 10,000 kb window) using the 1000 Genomes Project European ancestry reference panel via PLINK software. We extracted the effect estimates (beta) and standard errors of SNPs strongly associated with the exposure from exposure GWAS (thyroid function traits) and extracted the same SNPs from the outcome GWAS (ALS). The data were then harmonized to express the effect sizes on the same allele and remove palindromic SNPs (28), see supplementary material 1.

### Main MR analyses

Our main analysis focused on investigating the causal effect of genetically predicted thyroid function traits on amyotrophic lateral sclerosis (ALS), with the inverse variance weighting (IVW) as our primary analytical method. The IVW provides a causal effect estimate by performing a weighted regression of SNP-outcome associations on SNP-exposure associations, with the intercept fixed at zero. All analyses were performed using R Studio version 4.4.2 and the MRCIEU TwoSampleMR R package.

### Bidirectional MR analyses

Bidirectional two-sample MR was also performed to check for reverse causality and to examine the effects of genetic liability to ALS on thyroid function. All analyses were then conducted in the same manner as described above.

### Sensitivity analyses

We assessed the strength of the genetic instruments by estimating their F-statistic. By convention, if the F-statistic is greater than 10, MR causal effect estimates are unlikely to suffer from weak instrument bias that may violate the first MR assumption (23,28).

To assess the potential impact of directional horizontal pleiotropy on our overall causal estimates, we used MR-Egger regression, the weighted median, and the weighted mode methods. The Inverse Variance Weighting (IVW) method assumes zero horizontal pleiotropy, whereas MR-Egger regression incorporates an intercept term to identify and adjust for pleiotropic biases, regardless of the genetic instrument’s validity (29). The weighted median method can accommodate up to 50% of invalid instruments, while the weighted mode method can handle more than 50% of invalid instruments (29,30). We also applied the Mendelian Randomization Pleiotropy Residual Sum and Outlier (MR-PRESSO) method, which detects and corrects for overall pleiotropic outliers via three main steps: the global test, outlier detection, and the distortion tests, as elaborated upon elsewhere (31). Comparable effect estimates across these methods may suggest an actual causal effect.

Cochran’s Q test, was also applied to assess for the presence of heterogeneity among the estimates, with P-value > 0.05 assumed to indicate no significant heterogeneity (28). In cases where significant substantial heterogeneity was detected, a leave-one-out analysis was conducted to identify potential outliers with large effects by sequentially re-estimating the total effect after excluding one SNP at a time.

## RESULTS

### Genetic Instrument strength

We used the F-statistic to assess the strength of our genetic variants. The average was 80, with values ranging from 29 to 2,183. These results suggest that weak instrument bias is unlikely in our study and support the first MR assumption: that the genetic variants are strongly associated with the exposure.

### Causal effects of main analysis

We found evidence suggesting that genetic predisposition to autoimmune hyperthyroidism may be associated with 4% reduction in the odds of ALS (IVW odds ratio = 0.96; 95% CI 0.93 – 0.99, P = 0.03) (Figure 2a), with weighted median and weighted mode estimates directionally consistent with IVW estimates (Figures 2a and 3a). However, no evidence supported a causal relationship between other genetically predicted thyroid function traits and ALS risk, including TSH (IVW odds ratio = 0.99; 95% CI: 0.94 – 1.04; P = 0.71), FT4 levels (IVW odds ratio = 1.03; 95% CI: 0.96 – 1.12; P = 0.36), and autoimmune hypothyroidism (IVW odds ratio = 1.01; 95% CI: 0.98 – 1.05; P = 0.39). These findings are presented in Figures 2a and 3a.

**Figure 2.**
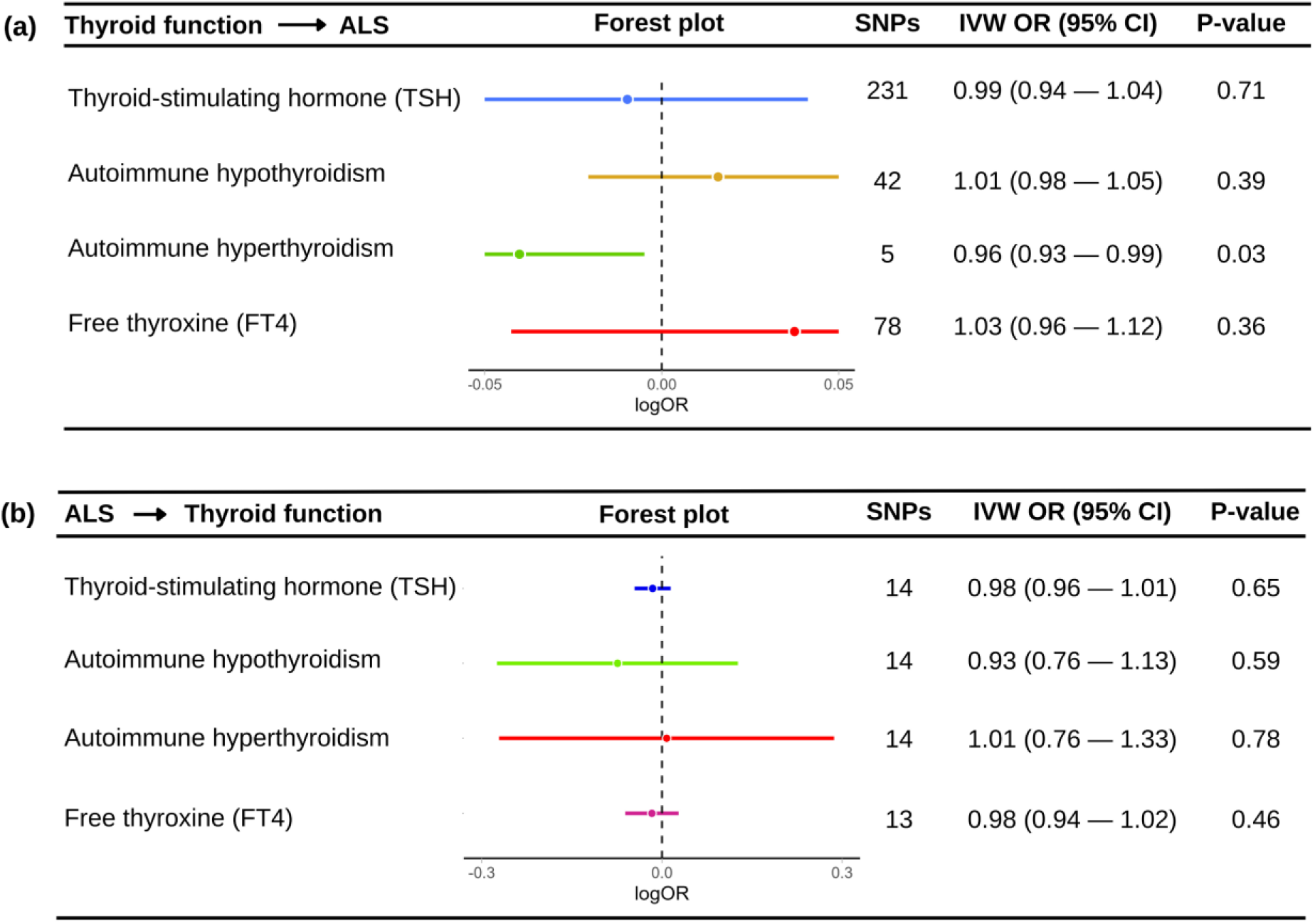
Inverse variance weighted (IVW) effect estimates. (a) genetically predicted thyroid function traits vs amyotrophic lateral sclerosis (ALS). (b) genetic liability to ALS vs thyroid function traits. Causal effect estimates are presented per log odds ratio increase of the outcome trait, with 95% confidence intervals.

**Figure 3.**
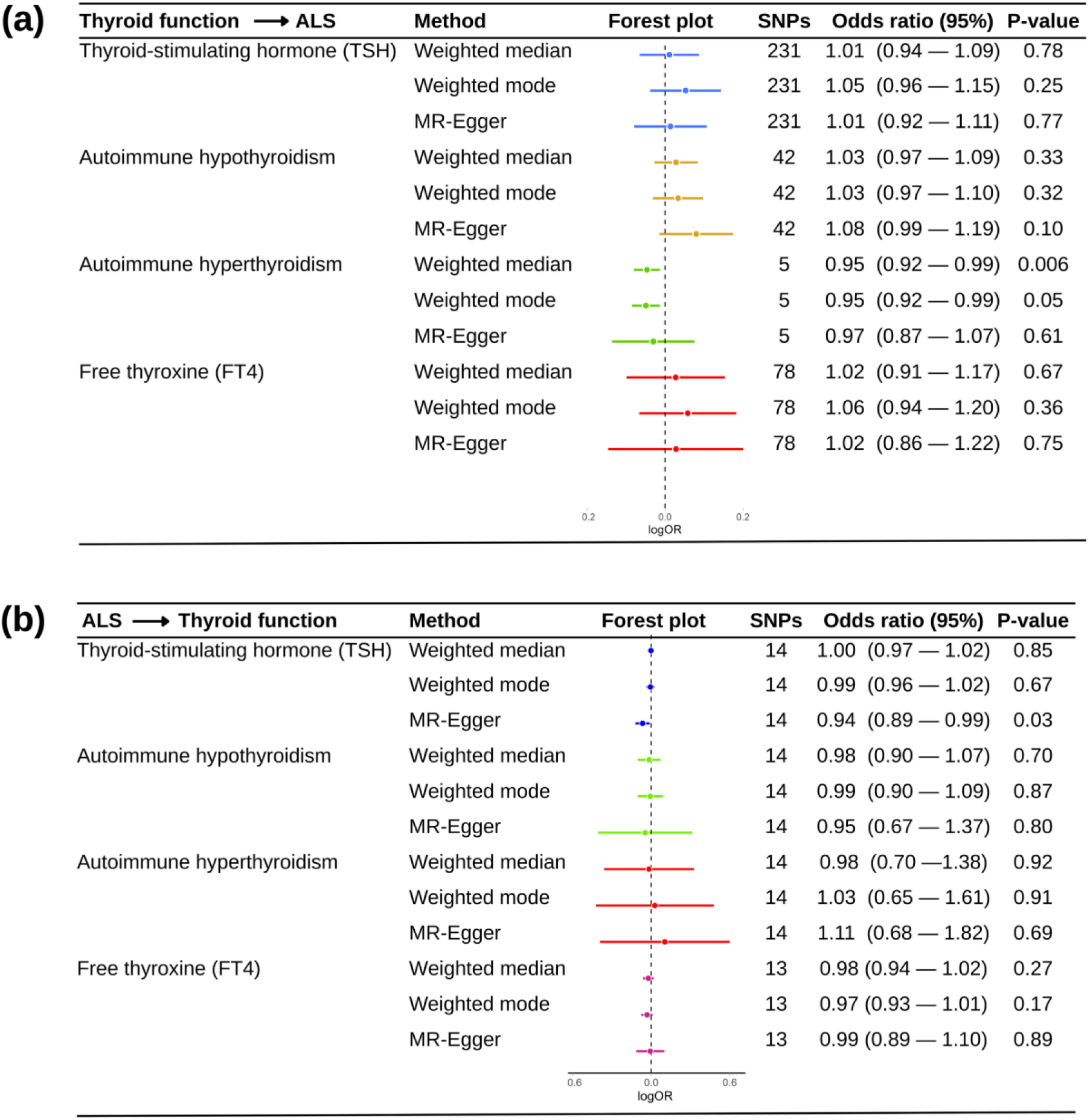
(a) Weighted median, weighted mode, and MR-Egger effect estimates of genetically predicted thyroid function traits on amyotrophic lateral sclerosis (ALS) using two-sample Mendelian randomization. (b) Bidirectional Mendelian randomization analysis showing the weighted median, weighted mode, and MR- Egger effects of genetic liability to ALS on thyroid function traits. Causal effect estimates are presented per log odds ratio increase of the outcome trait, with 95% confidence intervals.

### Causal effects of bidirectional analysis

In the bidirectional analysis, across IVW method, we found no evidence of a causal association between genetic liability to amyotrophic lateral sclerosis (ALS) and genetically predicted thyroid function traits, including TSH (IVW odds ratio = 0.98; 95% CI: 0.96 – 1.01; P = 0.65), FT4 levels (IVW odds ratio = 0.98; 95% CI: 0.94 – 1.02; P = 0.46), autoimmune hypothyroidism (IVW odds ratio = 0.93; 95% CI: 0.76 – 1.13; P = 0.59), and autoimmune hyperthyroidism (IVW odds ratio = 1.01; 95% CI: 0.76 – 1.33; P = 0.78). However, MR-Egger estimates suggest that genetic liability to ALS may lower TSH levels (MR-Egger odds ratio = 0.94; 95% CI: 0.89 – 0.99; P = 0.03). These findings are illustrated in Figures 2b and 3b.

### Evidence of heterogeneity and pleiotropy

Across both the main MR and bidirectional analyses, evidence of instrument heterogeneity was observed in most cases, except in the analysis examining the genetic liability to autoimmune hyperthyroidism vs ALS (Q = 6.4, P = 0.09), and autoimmune hypothyroidism vs ALS (Q = 43.3, P = 0.33), where Cochran’s Q tests showed no strong evidence of heterogeneity (Table 2). Similarly, for the analysis of genetic liability to ALS on autoimmune hyperthyroidism, Cochran’s Q test indicated no strong evidence of heterogeneity (Q = 16.8, P = 0.16; Table 2). Overall, leave-one-out analyses did not identify any single SNP as disproportionately influential, suggesting minimal variation in individual SNP effects (Figure 4).

**Figure 4.**
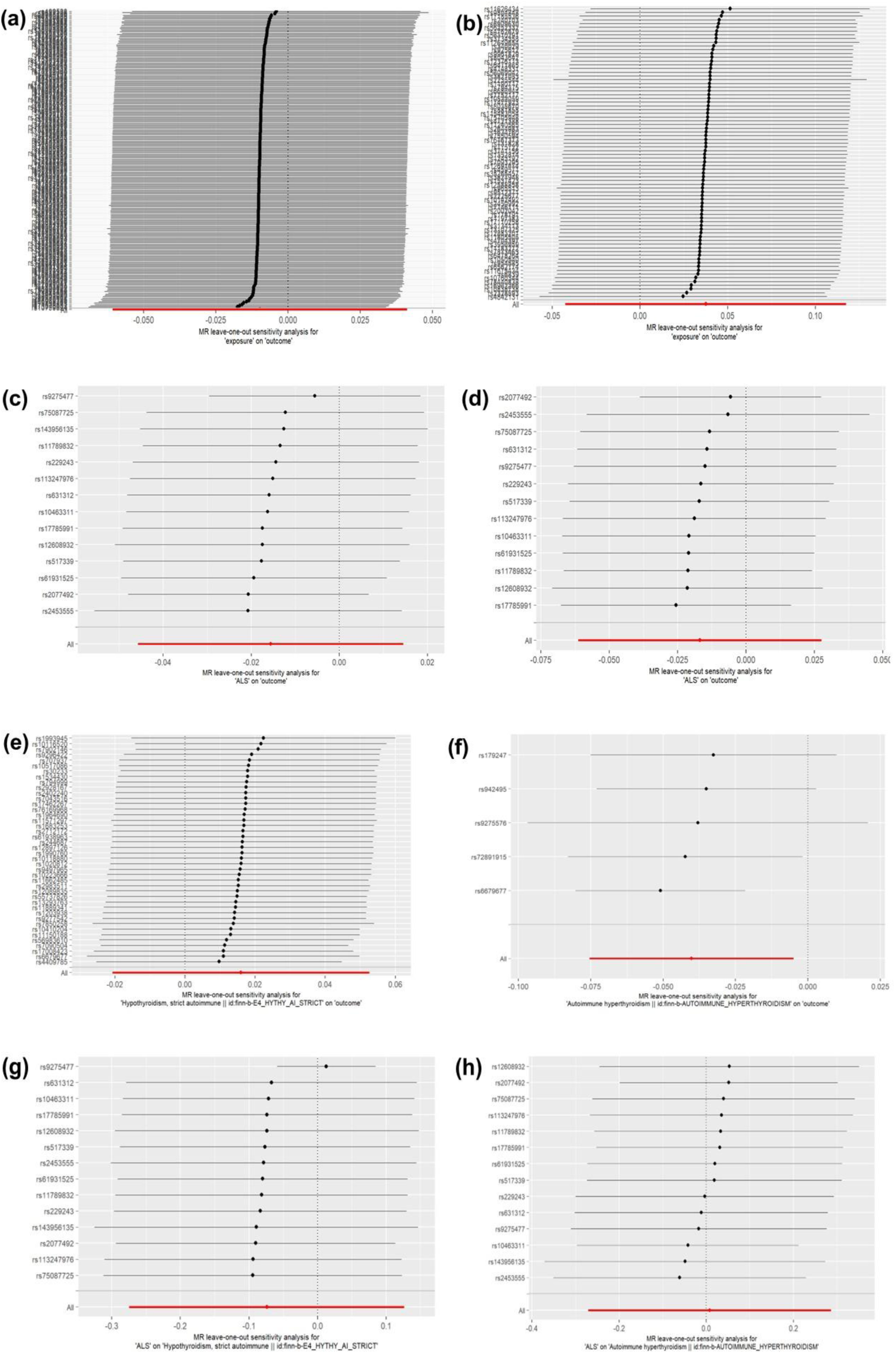
Leave-one-out analysis of: (a) TSH vs ALS, (b) FT4 vs ALS, (c) ALS vs TSH, (d) ALS vs FT4, (e) Autoimmune hypothyroidism vs ALS, (f) Autoimmune hyperthyroidism vs ALS, (g) ALS vs autoimmune hypothyroidism, (f) ALS vs autoimmune hyperthyroidism.

**Table 2:** Heterogeneity and pleiotropy test results for MR and bidirectional MR analyses.

| Exposure | Outcome | Heterogeneity |  | Pleiotropy |  |  |  |
| --- | --- | --- | --- | --- | --- | --- | --- |
|  |  | MR-Egger |  | MR-Egger intercept |  | MR-PRESSO global test |  |
|  |  | Q | P-value | Intercept | P-value | RSS obs | P-value |
| TSH | ALS | 307 | 0.0004 | -0.0012 | 0.55 | 311 | 0.001 |
| Autoimmune Hypothyroidism | ALS | 43 | 0.33 | -0.0079 | 0.15 | 55.97 | 0.20 |
| Autoimmune hyperthyroidism | ALS | 6.4 | 0.09 | -0.0061 | 0.85 | 8.47 | 0.31 |
| FT4 | ALS | 108 | 0.009 | 0.00046 | 0.90 | 311 | 0.0012 |
| ALS | TSH | 27.1 | 0.007 | 0.007 | 0.06 | 41.69 | 0.002 |
| ALS | Autoimmune Hypothyroidism | 140 | <<0.005 | -0.0045 | 0.86 | 159 | <0.001 |
| ALS | Autoimmune hyperthyroidism | 16.8 | 0.16 | -0.016 | 0.65 | 19.39 | 0.22 |
| ALS | FT4 | 40 | 0.001 | -0.0011 | 0.85 | 41.69 | 0.001 |

The Egger-intercepts for our instruments suggested minimal evidence of horizontal pleiotropy (Table 2). Although significant MR-PRESSO global test findings were observed in some instances (Table 2), none of the analyses with significant global tests also exhibited significant outlier and distortion tests to suggest a pleiotropic bias. Overall, causal estimates continued to align directionally with findings from other MR methods, even after outlier adjustments (see supplementary material 2).

## DISCUSSION

### Brief summary of main findings

In this study, we investigated whether genetically predicted thyroid function traits are linked with ALS using a two-sample Mendelian randomization approach. We found modest evidence of a negative causal effect between genetically predicted autoimmune hyperthyroidism and ALS. We also investigated the bidirectional causal effect of genetic liability to ALS on thyroid function, where we found limited evidence of a negative association with genetically predicted TSH and FT4 levels.

### Comparison to previous studies

To the best of our knowledge, this is the first study to explore genomic data to examine the causality of hyperthyroidism on ALS, and among the few that have employed other epidemiological approaches to investigate this association. Our observation of a potentially protective effect of genetically predicted autoimmune hyperthyroidism appears to contrast with studies that found no overall abnormalities in thyroid hormone levels in ALS patients (18,32–34) While a few studies (34,35) have observed the coexistence of hyperthyroidism and hypothyroidism among ALS patients, the directionality and clinical significance of this association remain unclear and warrant further rigorous investigation. In our bidirectional analysis, we found limited and method-specific evidence that genetic liability to ALS may be associated with lower levels of TSH. Specifically, this association was observed only using the MR Egger method, while other approaches, including IVW, Weighted Median, and Weighted Mode, did not support this finding. This suggests that the observed association may be driven by pleiotropy or other biases rather than a true causal effect. Nonetheless, our result partially aligns with prior studies reporting a higher prevalence of hypothyroidism among individuals with ALS (36).

These earlier studies, while valuable, relied primarily on observational designs, which may have limited their ability to account for confounding and detect directional associations. While our MR study provides modest evidence for a potential causal effect of genetically predicted autoimmune hyperthyroidism on reducing ALS risk, it is crucial to interpret this finding within the framework of MR. Our analysis infers the effect of the lifelong genetic predisposition to altered thyroid function, as proxied by the selected genetic variants, on ALS risk. However, the precise biological mechanisms through which this genetic predisposition influences ALS risk remain to be elucidated and warrant further investigation.

While the clinical significance of this our observed effect is unclear, it warrants careful consideration, the small magnitude of the effect suggests that genetically predicted autoimmune hyperthyroidism is unlikely to be a major determinant of ALS risk at the individual level. Thus, our result does not imply that inducing hyperthyroidism would be a clinically viable strategy due to the inherent risks of thyroid hormone imbalances and other associated risks. Nevertheless, we may infer that certain immune or hormonal pathways involved in autoimmune hyperthyroidism might confer some neuroprotective effects, and understanding these pathways is essential for translating this genetic association into potential clinical insights.

### Strengths and limitations

This study has several strengths. First, the use of a Mendelian Randomization (MR) approach allowed us to infer causality while minimizing confounding and reverse causation. Second, the use independent, non-overlapping samples of the same ancestry for exposure and outcome. Additionally, sensitivity analyses including MR-Egger, weighted median, weighted mode, leave-one-out, and MR-PRESSO revealed minimal heterogeneity and pleiotropy, confirming the robustness of our findings. Finally, the strong genetic instruments used for our analysis as indicated by the F-statistic

However, limitations must be acknowledged. Our analyses were restricted to individuals of European ancestry, potentially limiting generalizability to populations with diverse genetic and environmental backgrounds. Furthermore, reliance on summary-level data restricted our ability to explore nonlinear effects or conduct stratified analyses, especially considering that ALS is twice as prevalent in male compared to females. Finally, our study focused on common genetic variants, and likely underrepresented rare variants with larger effects.

### Future directions

Despite the limitations in this study, our findings contribute original insights to an understudied area in neuroepidemiology and have several implications for further research into the role of thyroid function in neurodegenerative disease risk. First, while this study did not specifically aim to examine the relationship between Graves’ disease and ALS, it is noteworthy that autoimmune hyperthyroidism, often caused by Graves’ disease, may share underlying mechanisms with ALS, given our finding of a protective effect of hyperthyroidism on ALS risk. Thus, future research should explore this potential overlap to elucidate the role of autoimmune thyroid disorders in neurodegenerative diseases. Additionally, studies should include diverse ethnic populations to assess whether genetic variants associated with ALS, which were not significant in European populations, exhibit associations in other groups. This approach would help identify population-specific genetic factors and enhance the generalizability of findings across varied genetic and environmental contexts. Furthermore, incorporating whole-exome sequencing (WES) and whole-genome sequencing (WGS) in future investigations could capture rare variants with large effects, potentially missed in this study. Finally, integrating these findings with multi-omics data (e.g. proteomics, metabolomics, and transcriptomics) would provide a more comprehensive understanding of ALS aetiology and progression, uncovering novel molecular mechanisms by capturing the complexity of the biological systems involved.

## CONCLUSION

This study advances our understanding of the causal relationship between thyroid function and ALS through the application of Mendelian Randomization (MR). We provide evidence that genetic predisposition to autoimmune hyperthyroidism may reduce ALS risk. As ALS remains an increasingly devastating global disease, these findings expand the knowledge of its genetic underpinnings and lay a foundation for future research into therapeutic strategies targeting thyroid function to mitigate ALS risk.

## Data Availability

The data used in this study were obtained from publicly available databases that adhere to ethical standards and legal requirements. Other data generated that support the findings of this study are available upon request from the corresponding author.

https://gwas.mrcieu.ac.uk/

https://ebi.ac.uk/gwas/

## ACKNOWLEDGEMENTS

We acknowledge and appreciate the contributions of the researchers and participants whose data were used in this study, particularly from the GWAS datasets made available by the FinnGen consortium, the ThyroidOmics consortium, and the UK Biobank.

